# Classification of colon cancer patients into Consensus Molecular Subtypes using Support Vector Machines

**DOI:** 10.1101/2023.05.22.23290335

**Authors:** Necla Kochan, Barıs Emre Dayanc

**Author notes:** Corresponding author at: Basic Medical Sciences, Faculty of Medicine, Izmir University of Economics, Balcova, Izmir, Turkey. *E-mail address:* (B.E. Dayanc).

## Abstract

**Objective:** The molecular heterogeneity of colon cancer has made classification of tumors a requirement for effective treatment. One of the approaches for molecular subtyping of colon cancer patients is the Consensus Molecular Subtypes (CMS) developed by the Colorectal Cancer Subtyping Consortium (CRCSC). CMS-specific RNA-Seq dependent classification approaches are recent with relatively low sensitivity and specificity. In this study, we aimed to classify patients into CMS groups using RNA-seq profiles.

**Methods:** We first identified subtype specific and survival associated genes using Fuzzy C-Means (FCM) algorithm and log-rank test. Then we classified patients using Support Vector Machines with Backward Elimination methodology.

**Results:** We optimized RNA-seq based classification using 25 genes with minimum classification error rate. Here we report the classification performance using precision, sensitivity, specificity, false discovery rate and balanced accuracy metrics.

**Conclusion:** We present the gene list for colon cancer classification with minimum classification error rates. We observed the lowest sensitivity but highest specificity with CMS3-associated genes, which is significant due to low number of patients in the clinic for this group.

## 1 Introduction

Colon cancer is one of the most common cancer types worldwide and the second and third leading cause of cancer deaths for men and women, respectively. Approximately 8% of cancer-related deaths in the world are associated with colon cancer (Schweiger et al., 2013). The molecular heterogeneity and complexity of colon cancer makes the prediction of disease and potential treatments harder. In order to better characterize and resolve the heterogeneity, researchers have focused on sub-typing of colon tumors. The Colorectal Cancer Subtyping Consortium (CRCSC) published a study where CRC patients were stratified into four distinct Consensus Molecular Subtypes (CMS) and one unknown group, in which patients had no CMS information (nolabel) (Guinney et al., 2015). For CMS subtyping, CRCSC-developed *CMSclassifier* algorithm which uses hundreds of genes from the whole genome data (Buechler et al., 2020). Then an R package called *CMScaller* was developed and published by (Eide et al., 2017), which uses more than five hundred genes from the whole genome data. However, until Buechler *et al*. recently published RNA-Seq and microarray data-based CMS subtyping (*ColoType*) with 40 genes (Buechler et al., 2020), there was no RNA-Seq based CMS specific approach. The rationale of our study is that microarray and RNA sequencing technologies are inherently different and both technologies have some shortcomings as summarized in (Eilertsen et al., 2020), such as inherent technical biases observed with microarrays related to cross hybridization and a limited dynamic range of expression (Wang et al., 2009). These would have an impact on subtype distributions according to clinically relevant classification frameworks such as CMS. As long as the systematic biases are addressed (representation of short genes and genes with low expression levels), RNA-Seq represents a reliable and preferred method for transcriptomic subtyping of colon cancer by whole transcriptome profiling (Wang et al., 2009).

The studies show that cancer classification based on gene expression data has become an important part of modern medicine. Therefore, in this study, we applied Support Vector Machines (SVMs) to classify CRC patients on CMS status based on their gene expression levels. We particularly focused on RNA-Seq data that includes CMS information for each patient.

## 2 Materials and Methods

### 2.1 Gene Expression Data and Survival Data

RNA-Seq data for CRC patients is obtained from the TCGA database using **TCGAbiolinks** package. We then considered patients with Primary Tumor (PT) and Solid Tissue Normal (STN). Among these patients, we selected the patients who were diagnosed with primary adenocarcinoma but received no therapy. We used Disease Specific Survival (DSS) data for the survival analysis and the survival data of TCGA COAD samples is obtained from (Liu et al., 2018). In order to identify the molecular subtype specific prognostic genes in colon cancer, we downloaded used subtype information of TCGA COAD patients from synapse.org. After gathering all the information we require, we had 29 patients in CMS1, 82 patients in CMS2, 27 patients in CMS3 and 58 patients in CMS4. We finally filtered the genes, which have very low or no expression using FPKM (Fragments Per Kilobase of transcript per Million) values. We filtered the genes whose FPKM values are below 0.5 in both PT and STN to avoid the systematic bias of RNA-Seq data on genes with low expression. After filtering, we had 14,334 genes left for further analysis.

### 2.2 Identification of subtype specific prognostic genes for colon cancer using FCM

In order to identify subtype specific prognostic genes, analyses are performed for each CRC molecular subtype, separately. Then the FCM clustering algorithm is applied to stratify patients into two clusters (groups) with membership degrees for each patient and cluster centers. The algorithm assigns each patient to the one of the clusters with the maximum membership degree, which displays the degree of belonging to the corresponding cluster. A representative FCM clustering of *FOXJ1* gene for each subtype was depicted as violin plot as in Figure 1.

**Figure 1.**
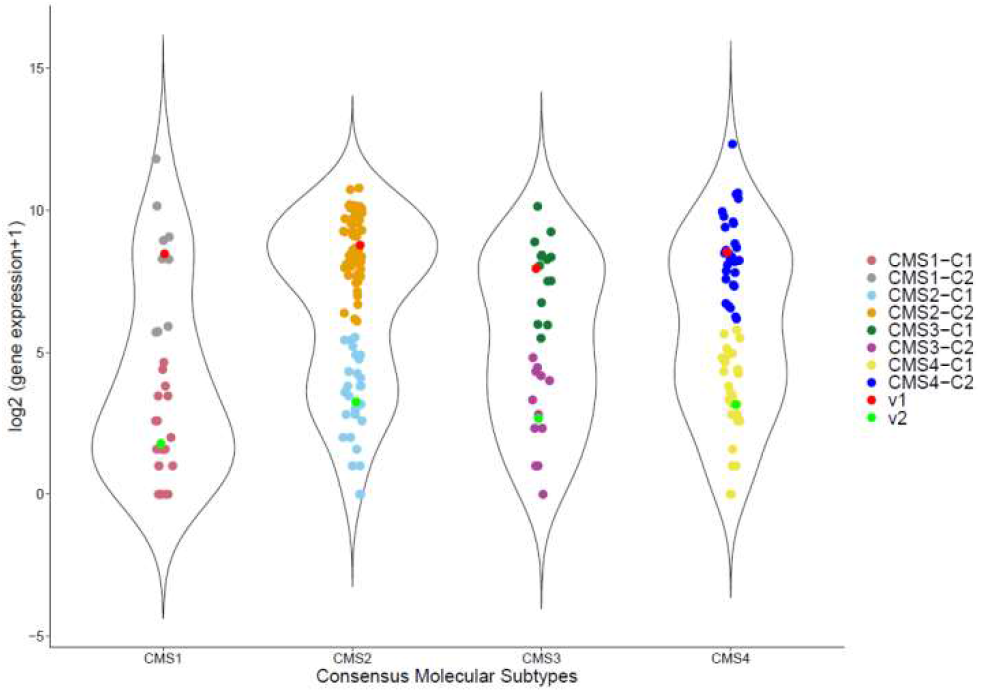
FCM clustering. Stratification using FCM where C1 and C2 are cluster centers for cluster I and II, respectively. The points in the same cluster are similar. The points that overlap are assigned to one of the clusters with respect to the maximum membership degrees.

Then the genes that can significantly differentiate between the survivals of these two groups are chosen with a cutoff *p* value of 0.01. We note here that log2 expression values are used. By applying FCM based approach we obtained 86 genes for CMS1, 148 genes for CMS2, 8 genes for CMS3 and 53 genes for CMS4 that are statistically significant (*p<0*.*01*). After reducing the number of genes, we performed univariate Cox regression to get the most informative genes for further analysis. As a result of univariate Cox regression, these numbers are reduced to 6, 48, 2 and 25 for CMS1, CMS2, CMS3 and CMS4, respectively. We note here that since we could not find any significant genes for CMS3 with a maximum p-value of 0.01, we have taken *p*-value cut-off as 0.05 for CMS3 group to have at least one gene for each molecular subtype.

### 2.3 Gene Selection

With the advent of next generation sequencing, one can detect tens of thousands of genes simultaneously and this provides a deep insight into cancer classification problems. The major challenge in gene expression data classification is to extract disease/cancer related information from the huge amount of data, which consists of redundant information and noise. Therefore, obtaining the significant amount of information is a key step for gene expression data classification problems.

Instead of starting with more than 20,000 genes and applying any feature selection methods, we started with the molecular subtype specific prognostic genes, which are identified in the previous section. We claim that these genes have crucial roles in colorectal cancer subtype classification. Then we searched for the gene list using a backward elimination method (Figure 2).

**Figure 2.**
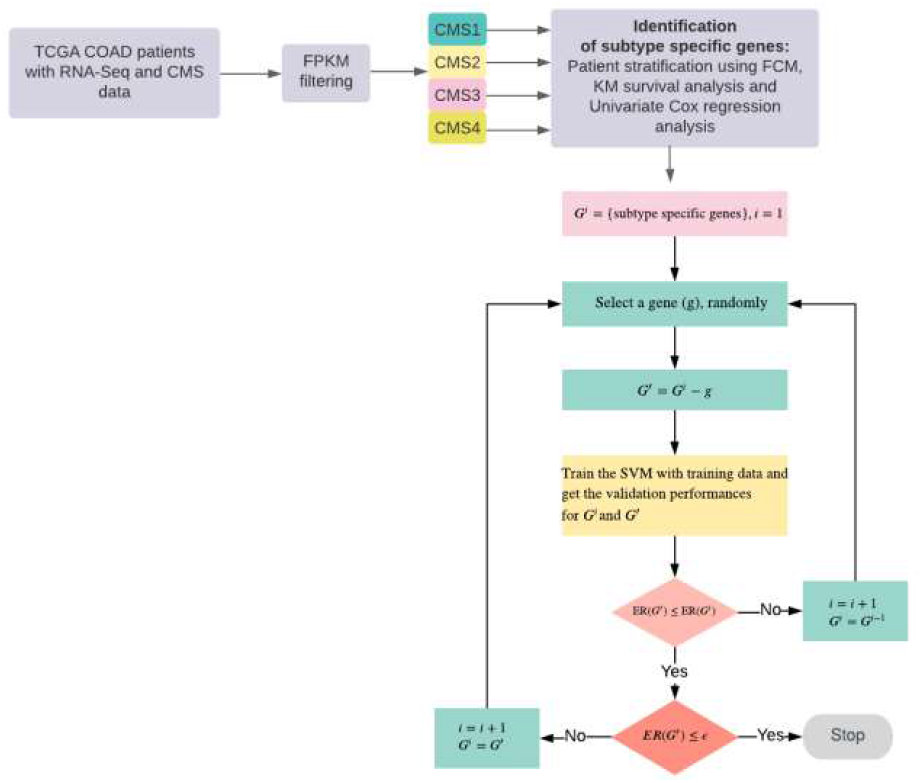
Flowchart of our method. Identification of prognostic genes and backward elimination algorithm for gene selection in order to classify CRC patients. ER: Error Rate, *G*: the set of genes

**Figure 3.**
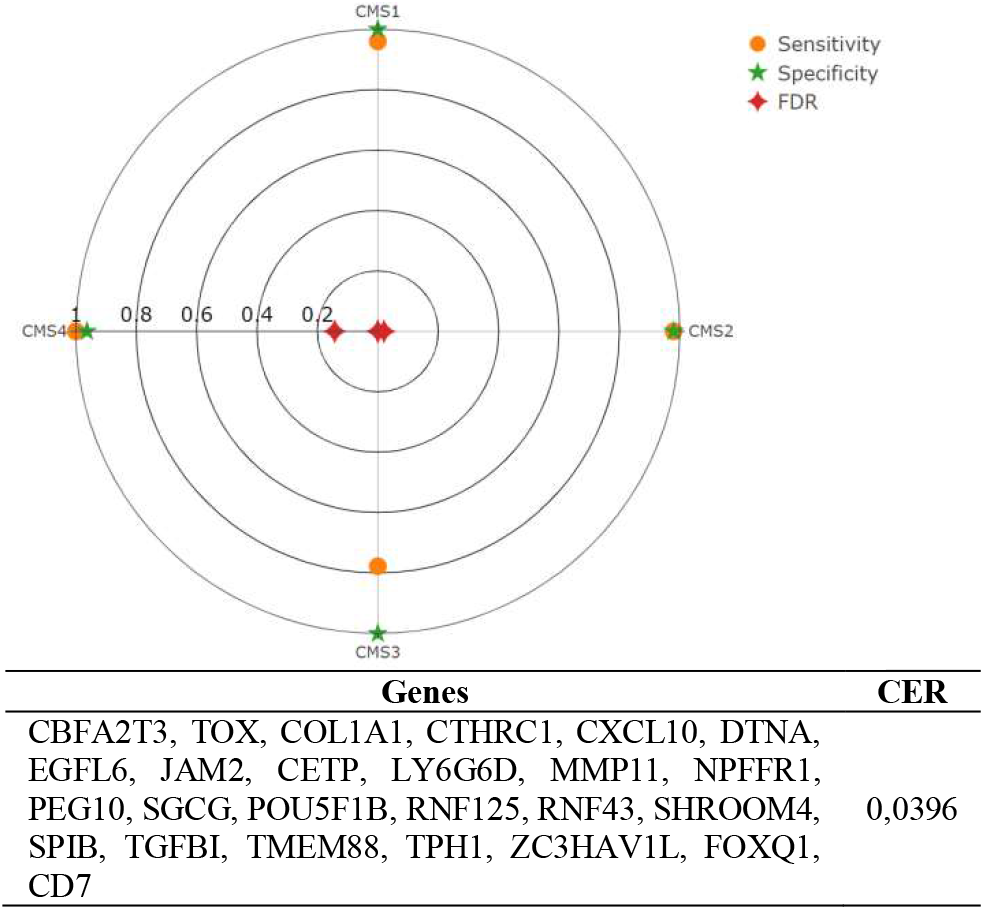
Performance analysis using 25 genes. Classification performances of each CMS in terms of sensitivity, specificity and False Discovery Rate (FDR). Overall performance is given in terms of classification error rate, which is the ratio of misclassified patients over all patients in the test set.

### 2.4 SVM Classification

Support Vector Machines (SVMs), which was developed by (Vapnik, 2000) is a kernel-based machine-learning algorithm. It has been applied to numerous areas such as pattern recognition, medicine, bioinformatics, biological and other sciences.

SVM finds an ideal decision boundary, which is also called ideal separation hyperplane in order to separate classes. The ideal decision boundary or hyperplane is determined according to the maximum margin principle. Briefly, the algorithm finds the ideal hyperplane that maximizes the distance between classes. The vectors which define these hyperplanes are called support vectors.

If the classes are linearly separable then SVM performs efficiently and splits the classes without an overlap. However, a perfect separation may not be observed in many real-life data sets. In that case, SVM searches for the hyperplane, which minimizes the classification error rate and maximizes the margin (Bishop, 2006). If the data is linearly non-separable, SVM uses kernel functions, i.e. linear, nonlinear sigmoid and radial basis kernel in order to convert the non-separable data into linearly separable data form.

#### Classification

Cancer classification on the basis of gene expression data has become an important part of modern medicine as it provides an objective and accurate diagnosis of different types of cancers/diseases. A number of machine learning approaches e.g., Support Vector Machines (SVMs), Random Forest (RF) and *k*-Nearest Neighbour (*k*NN) have been applied on gene expression data classification. On the other hand, it is a challenging problem since patients who have cancer is not classified with their gene expression levels but tumor pathological information. This shows that there is a gap in the literature in terms of cancer classification at gene expression level. Colon cancer is one of those cancer types that need further investigation. Therefore, in this study, we applied the SVM algorithm for the colorectal cancer patient classification, as it is one of the most powerful supervised learning algorithms. SVM is performed using the “**e1071**” package in R with a radial basis kernel.

### 2.5 Performance Evaluation Metrics

The class-specific performances were calculated according to the precision, sensitivity, specificity, false discovery rate (FDR) and balanced accuracy, which are defined as follows:

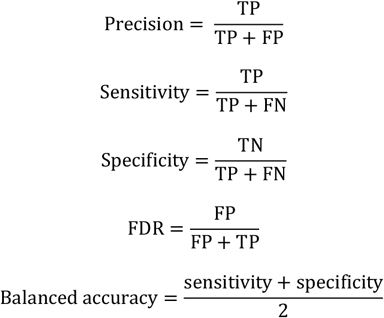

where TP represents True Positives, FP represents False Positives, TN is True Negatives and FN is False Negatives. Overall performance is measured by the Classification Error Rate (CER), which is defined as follows:

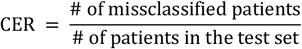

### 2.6 Statistical Analysis

Statistical analyses were performed using R language (v.4.0.2). Kaplan-Meier and log-rank tests were performed to assess survival differences between clusters and risk groups. Univariate Cox regression analysis was performed using “survival” and “survminer” packages in R. *p* values below 0.01 were considered statistically significant for all comparisons except for CMS3 subtype specific prognostic genes, as described before.

## 3 Results

We considered the discovery set used in identifying the subtype specific prognostic genes as the training set and the test set is the TCGA COAD data that are not included in the training set. The CMS clinical information of all patients (training and test sets) was downloaded from “synapse.org”. The training set (66% of the dataset) is used to train the SVM classification model and the test set (34% of the dataset) is used to measure the classification error rate.

The results showed that using 25 genes, we reached the minimum classification error rate which is 0,0396 (Figure 2). Moreover, the subtype specific statistics are given in terms of precision, specificity, sensitivity, FDR, balanced accuracy (Table 2). We see that CMS3 has the smallest sensitivity but the highest specificity and the other subtypes (CMS1, CMS2 and CMS4) have not only high sensitivity but also high specificity for the 25-gene list.

**Table 1.** Data description. The number of patients used for the training dataset and test dataset, accordingly.

|  | CMS1 | CMS2 | CMS3 | CMS4 | Total |
| --- | --- | --- | --- | --- | --- |
| Training Set | 28 | 82 | 27 | 58 | 195 |
| Test Set | 125 | 49 | 9 | 18 | 101 |
| Total | 53 | 131 | 36 | 76 | 296 |

**Table 2.** Classification performance. 25 genes are used which leads to minimum classification error rate

|  | Precision | Sensitivity | Specificity | FDR | Balanced accuracy |
| --- | --- | --- | --- | --- | --- |
| CMS1 | 1,0000 | 0,9600 | 1,0000 | 0,0000 | 0,9800 |
| CMS2 | 0,9796 | 0,9796 | 0,9800 | 0,0204 | 0,9798 |
| CMS3 | 1,0000 | 0,7778 | 1,0000 | 0,0000 | 0,8889 |
| CMS4 | 0,8571 | 1,0000 | 0,9634 | 0,1429 | 0,9817 |

## 4 Discussion

In this study, we have discovered two gene lists for colon cancer classification with minimum classification error rates. SVM is a kernel-based algorithm and one of the widely applied classification algorithms in bioinformatics due to its high accuracy (Zhi et al., 2018). To the best of our knowledge, this is the first study to classify TCGA COAD patients using a new pipeline, which involves identifying survival-associated genes and applying SVMs with backward elimination. By utilizing this novel method, we aim to improve the accuracy of classification and identify potential prognostic biomarkers for colon cancer.

Consensus Molecular Subtypes of colorectal cancer patients are determined by molecular tumor pathologic information. Although patient treatment modalities for colon cancer today is prescribed by tumor staging, very few tools could be used to guide clinical decisions until recently. Buechler group (Buechler et al., 2020) developed the 40-gene ColoType risk score model for CRC patient classification with an 88% performance in TCGA-COAD RNA-Seq data. In order to compare our results with this study, we used the 40 genes reported in their study in our training and test sets using SVM classification algorithm. The error rate was measured as 0.13 when we used 40 genes with our test set.

It is important to note that this study is specific to CRC RNA-Seq data with additional CMS information, i.e., patients without CMS information are excluded. Thus, our approach is limited only to publicly available TCGA COAD data. We tested a single cohort, therefore other cohorts with CMS information can be further investigated.

In order to identify CMS-specific genes, we considered the patients who had primary adenomacarcinoma and received no treatments. We selected the patients who had DSS clinical information provided given in (Liu et al., 2018) that DSS is more specific than Overall Survival. Due to the low number of CMS3 patients in TCGA COAD study and as our patient selection criteria further limited the number of patients in each molecular subtype, patient number in CMS3 is low. The reliability of this study could further be improved with more RNA-Seq gene expression data and patients’ CMS subtype information together.

## Data Availability

All data produced in the present study are available upon reasonable request to the authors

